# Mendelian randomization analyses show that higher acetyl-carnitine and carnitine levels in blood protect against severe Covid19

**DOI:** 10.1101/2021.05.31.21257910

**Authors:** Nabila Kazmi, George Davey Smith, Sarah J Lewis

**Affiliations:** MRC Integrative Epidemiology Unit (IEU), Bristol Medical School, University of Bristol, Bristol, UK; Population Health Sciences, Bristol Medical School, University of Bristol, Bristol, UK

**Keywords:** Covid19, carnitine, acetyl-carnitine, Mendelian randomization

## Abstract

**Background:** Severe Covid19 is characterised by a hyperactive immune response. Carnitine, an essential nutrient, and it’s derivative acetyl-carnitine can downregulate proinflammatory cytokines and has been suggested as a potential treatment for the disease.

**Methods:** We carried out Mendelian randomization analyses using publicly available data from a large genome wide association study (GWAS) of metabolites and a collaborative genome wide study of Covid19 to investigate the nature of the relationship between carnitine and acetyl-carnitine and Covid19 infection, hospitalisation with Covid19 and very severe Covid19. We used the same methodology to determine whether carnitine was associated with co-morbidities commonly found among those with severe Covid19.

**Results:** We found evidence of a protective effect against very severe Covid19 for both carnitine and acetyl-carnitine, with around a 40% reduction in risk associated with a doubling of carnitine or acetyl-carnitine (carnitine odds ratio (OR) = 0.56, 95% confidence intervals (CI) 0.33 to 0.95, p=0.03 and acetyl-carnitine OR=0.60, 95% CI 0.35 to 1.02, p=0.06), and evidence of protective effects on hopitalisation with Covid19. For acetyl-carnitine the largest protective effect was seen in the comparison between those hospitalised with Covid19 and those infected but not hospitalised (OR=0.34, 95%CI 0.18 to 0.62, p=0.0005).

**Conclusion:** Carnitine and acetyl-carnitine merit further investigation in respect to the prevention of severe Covid19.

## Introduction

The Covid19 pandemic in 2020/21 caused by the novel SARSCov2 virus has resulted in substantial loss of life and severe morbidity worldwide.^1^ Whilst the majority of people who contract the virus experience either no obvious symptoms or only mild symptoms and recover within 1-2 weeks, some individuals go on to develop severe disease and need hospitalisation, and a minority do not survive the illness.^2^ Even after controlling for age and other risk factors there is still substantial heterogeneity in response. Severe disease is characterised by a hyper-immune response or cytokine storm and those admitted to intensive care units have been shown to have high blood levels of inflammatory proteins.^3,4^ Treatment with dexamethasone and other corticosteroids which are anti-inflammatory drugs has been shown to be somewhat effective and are now routinely used to treat those with severe Covid19.^5^ Interleukin-6 inhibitors are also recommended as effective treatments,^6^ after the RECOVERY trial showed a benefit among those hospitalised for treatment for Covid-19.^7^

Acetyl-carnitine has also been hypothesised to be potentially effective against Covid19,^8^ leading to a small randomized controlled trial being initiated to study the effectiveness of this treatment in hospitalised Covid19 cases.^9^ Acetyl-carnitine is the acetylated form of carnitine, an essential nutrient which is ingested from eating meat or is made in small quantities in the body from amino acids lysine and methionine.^10^ Acetyl-carnitine is easily converted to carnitine in cells, by removal of the acetyl group by co-enzyme A, producing acetyl co-A (the primary substrate for Krebs cycle and production of ATP), thus the two are interchangeable and have an important biological function.^11^ However, acetyl-carnitine is more easily absorbed from the gut, and more readily crosses the blood-brain barrier and is therefore preferentially given as a supplement.^11^ Aside from playing a key role in fatty acid oxidation by transporting long chain fatty acyl-CoAs into the mitochondria for degradation by β-oxidation,^10,12^ carnitine has also been shown to downregulate pro-inflammatory cytokines including TNF-α, IL-6, and IL-1 in animal studies^13,14^ and it is for this reason it has been advanced as a potential agent which could prevent or treat the cytokine storm which occurs in severe Covid19 patients.

Large randomized controlled trials are expensive and time consuming and small trials may be underpowered to observe an effect. The ongoing randomized controlled trial of acetyl carnitine and Covid19 includes just 100 individuals and will not be definitive.^9^ Mendelian randomization studies, in which genetic variants associated with the exposure or treatment of interest are used as proxies for the exposure and genetically defined groups are compared for the outcome of interest, offer a cheap and quick way to prioritise potentially effective treatments for investigation in large randomized controlled trials.^15,16^ Mendelian randomization has already been used to show that genetic variants mimicking therapeutic inhibition of IL-6 receptor signalling protect against severe Covid19.^17^ We carried-out a two sample Mendelian randomization analysis of acetyl-carnitine and carnitine on Covid19 risk. We used publicly available data from a large genome wide association study (GWAS) of a metabolomics platform including carnitine and acetyl-carnitine^18^ for our genetic variant-exposure estimate and a large-scale collaborative genome wide study of Covid19 for our genetic variant-outcome estimate.^19^

## Methods

### Exposure instruments

We used single nucleotide polymorphisms (SNPs) as instruments in our Mendelian randomization analysis which have previously been identified as being associated with carnitine and acetyl-carnitine concentration measured in blood from 7,797 and 7,805 adults of European ancestry respectively.^18^ The study combined association results in TwinsUK and KORA (Cooperative Health Research in the Region of Augsburg) using inverse variance meta-analysis.^18^ Acetyl-carnitine and carnitine metabolites were profiled using liquid chromatography mass spectrometry (LC-MS). Two SNPs associated with acetyl-carnitine concentration (Supplemental material; Table S1) and seventeen SNPs associated with carnitine concentration (Supplemental material; Table S2) were identified at GWAS significance levels after removing those that were in linkage disequilibrium with the SNP having the smallest p-value in a given region. The variance in acetyl-carnitine and carnitine concentrations explained by the two and seventeen variants in our instruments were 1.9% and 12.4% respectively. The exposure-SNP associations were reported on a log base 10 scale, however we multiplied these estimates by 0.30103 to convert to a log base 2 scale and estimated the effect on our outcomes of interest per doubling of exposure. We also scaled our acetyl-carnitine exposure (by multiplying the betas and standard error from our inverse variance weighted analysis by log_10_(1.43)) to mimic the 43% increase in blood acetylcarnitine levels seen in a randomized controlled trial where acetyl-carnitine was given at 2g/day,^22^ to estimate what we might expect to observe with respect to the outcome of severe Covid19 when treating patients with mild symptoms as in the ongoing acetyl-carnitine trial for Covid19 (with the caveats that the age of participants in the two sets of trials is likely to be different and in our Mendelian randomization analysis of severe Covid19 we did not use those with known infection as the control group).^9^

### Outcome traits

GWAS results on COVID19 were obtained from the COVID-19 Host Genetics Initiative genome-wide association meta-analysis release 5, January 18, 2021 (without the 23andMe samples) performed in European populations.^19,21^ We included summary statistics from the following analyses as outcomes in our Mendelian randomization analyses 1) Covid19 versus population controls, comprising of 38984 cases and 1644784 controls 2) hospitalised with Covid19 versus population controls, comprising of 9986 cases and 1877672 controls 3) hospitalised with Covid19 versus not hospitalised with Covid19, 4829 cases and 11816 controls 4) severe Covid19 versus population controls, 5101 cases and 1383241 controls. More detail on case and control group definitions is provided in the text box 1 and in reference 20.

#### Box1 Covid19 outcomes and control groups used in this study

We carried-out four different analyses for this paper, the case definitions and control groups used for these are provided below:

**Any Covid19 -**all cases with reported SARS-CoV-2 infection with or without symptoms of any severity *versus* genetically ancestry matched samples without known SARS-CoV-2 infection *(test of likelihood of being infected)*

**Hospitalised with Covid19 versus population controls-**cases with moderate or severe COVID-19 defined as those hospitalised due to symptoms associated with the infection *versus* genetically ancestry matched samples without known SARS-CoV-2 infection *(test of disease severity)*

**Hospitalised with Covid19 versus Covid19 but not hospitalised-**cases with moderate or severe COVID-19 defined as those hospitalised due to symptoms associated with the infection *versus* people with confirmed Covid19 infection who had only mild disease or who were asymptomatic *(test of disease severity with more appropriate control group)*

**Severe Covid19-**critically ill COVID19 cases defined as those who required respiratory support in hospital or who were deceased due to the disease *versus* genetically ancestry matched samples without know SARS-CoV-2 infection *(test of disease severity)*

In sensitivity analyses, muscle weakness proxied by grip strength was used as a positive control outcome;^22^ we expected carnitine and acetyl-carnitine to show inverse associations with this trait as it has been shown to occur in those who have inborne errors of metabolism leading to carnitine deficiencies.^23^ We investigated the impact of carnitine and acetyl-carnitine on comorbidities that increase the risk of severe COVID19 to identify whether these may be on the causal pathway or to highlight any off target effects.^2^ Therefore, we included Alzheimer’s disease, breast cancer, chronic kidney disease, coronary heart disease, type II diabetes, prostate cancer and stroke in our MR analyses (details on these outcomes are provided in the supplemental material; Table S3). We also investigated the impact of carnitine and acetyl-carnitine on body mass index (BMI), fat mass, fat-free mass, height and C-reactive protein levels.

### Two sample MR analysis

MR analyses were performed using the inverse-variance weighted (IVW) method as our primary approach.^24^ For our carnitine instrument we were able to carry-out sensitivity analyses, but we were not able to for our acetyl-carnitine instrument because this only consisted of two SNPs. For our carnitine analyses we carried out weighted median^25^, weighted mode^26^, MR-Egger regression^27^ and MR-PRESSO analyses.^28^ We assessed for violations of the MR ‘no horizontal pleiotropy’ assumption using visual inspection of funnel,^29^ scatter, forest and leave-one-out plots and tests of heterogeneity between the SNPs.^30^ We tested whether our outcomes could affect carnitine and acetylcarnitine levels (reverse causation) using a Steiger test.^31^ Finally, we tested whether our analysis of carnitine and acetyl-carnitine and severe Covid19 were confounded by BMI, diabetes or coronary heart disease, and whether our analyses of carnitine and acetyl-carnitine levels on C-reactive protein levels were confounded by BMI by carrying out a multivariable Mendelian randomization (MVMR) analyses^32^.

In order to identify potential pleiotropic effects of the carnitine and acteylcarnitine SNPs used in our analysis we also searched for the effects of each genetic variant across all publicly available GWAS datasets using PheWAS (https://gwas.mrcieu.ac.uk/phewas/), a platform built specifically for this purpose.^33^We reported all phenotypes associated with our SNPs at significance level p<5×10^−8^ (supplemental material; Tables S1 and S2). For the acetyl-carnitine SNPs, because we were unable to run any sensitivity analyses, we assessed the similarity between the two SNPs for the traits which were found to be associated with either one at GWAS significant levels (supplemental material; Table S4), to determine whether they were likely to be a result of acetyl-carnitine levels or due to effects of the SNPs on other biological pathways.

Before proceeding with the analyses for our positive control and comorbidities we calculated the power to detect an odds ratio (OR) of 1.2 (or 0.80) per doubling of exposure for dichotomous outcomes with an alpha-level of 0.05, the variance explained in the exposure by the instrument and the sample size for comorbidities of Covid19, as described previously.^34^ A priori we planned to performed MR analyses on those outcomes that had power > 50%.

## Results

### Acetyl-carnitine and Covid19

We found modest evidence that a doubling in acetyl-carnitine concentration in blood reduces the risk of severe Covid19 by 40% (Table1; Figure 1 and supplemental material; Figures S1). Stronger evidence of a protective association was seen for hospitalisation with Covid19 versus Covid19 infection but not hospitalised where risk was reduced by 67%. We also found evidence of a protective effect of acetyl-carnitine on hospitalisation with Covid19 versus population controls (35% reduction in risk), although there was little evidence of any association of acetyl-carnitine with any Covid19 infection. When our analyses were scaled to detect an effect of a 43% increase in acetyl-carnitine, the level of increase expected from 2g per day supplementation as given in the ongoing trial,^9^ we found that risk of severe Covid19 decreased by 23% (OR: 0.77; 95% CI: 0.58 to 1.01). We were unable to run sensitivity analyses to test for pleiotropy for acetyl-carnitine as we only had 2 SNPs, but the two SNPs gave consistent estimates. One acetyl-carnitine SNP (rs274567) maps to *SLC22A5* which is a carnitine transporter gene, mutations of which lead to carnitine deficiency,^23^ The other SNP (rs1171614) maps to another solute carrier *SLC16A9*, Table S1 (supplemental material) shows traits which were associated with each SNP at GWAS significance level, these were blood cell characteristics, anthropometry, hand grip strength, inflammatory diseases, basal metabolic rate and other carnitines, all potentially downstream of acetyl-carnitine. Table S4 (supplemental material) shows the effect estimates, standard errors and p-values for both SNPs against each trait identified as being associated with either SNP at GWAS significance level. For all traits except urate/gout/allopurinol treatment effect estimates were in the same direction for both SNPs, and for all for all except granulocyte count, 2-tetradecenoyl carnitine, asthma and hand grip strength there was strong evidence of an association with both SNPs.

**Table 1.** Results from Mendelian inverse variance weighted analyses of carnitine and acetyl-carnitine on Covid19 and its subgroups

| Outcome | # SNPs | Odds ratio | LCI | UCI | P<sub>assoc</sub> | P<sub>het</sub> | P<sub>pltr</sub> |
| --- | --- | --- | --- | --- | --- | --- | --- |
| ***Acetyl-carnitine*** |  |  |  |  |  |  |  |
| Any Covid19 | 2 | 0.94 | 0.56 | 1.56 | 0.80 | 0.008 | - |
| Hospitalised with Covid19-A | 2 | 0.34 | 0.18 | 0.62 | 0.0005 | 0.94 | - |
| Hospitalised with Covid19-B | 2 | 0.65 | 0.42 | 1.00 | 0.05 | 0.23 | - |
| Severe Covid 19 | 2 | 0.60 | 0.35 | 1.02 | 0.06 | 0.79 | - |
| ***Carnitine*** |  |  |  |  |  |  |  |
| Any Covid19 | 17 | 0.81 | 0.68 | 0.98 | 0.03 | 0.58 | 0.38 |
| Hospitalised with Covid19-A | 17 | 0.82 | 0.44 | 1.53 | 0.52 | 0.34 | 0.03 |
| Hospitalised with Covid19-B | 17 | 0.80 | 0.56 | 1.15 | 0.23 | 0.77 | 0.09 |
| Severe Covid19 | 17 | 0.56 | 0.33 | 0.95 | 0.03 | 0.87 | 0.47 |

LCI = lower 95% confidence interval, UCI = upper 95% confidence interval, P<sub>assoc</sub> = P-value for association, P<sub>het</sub> = P-value for heterogeneity between instrumental variables, SNP causal estimates, P<sub>pltr</sub> = P-value for horizontal pleiotropy from MR-Egger intercept test; # SNPs = total number of SNPs used in the analysis, Hospitalised with Covid19-A = hospitalised with covid vs. not hospitalised with covid; Hospitalised with Covid19-B = hospitalised covid vs. population

**Figure 1.**
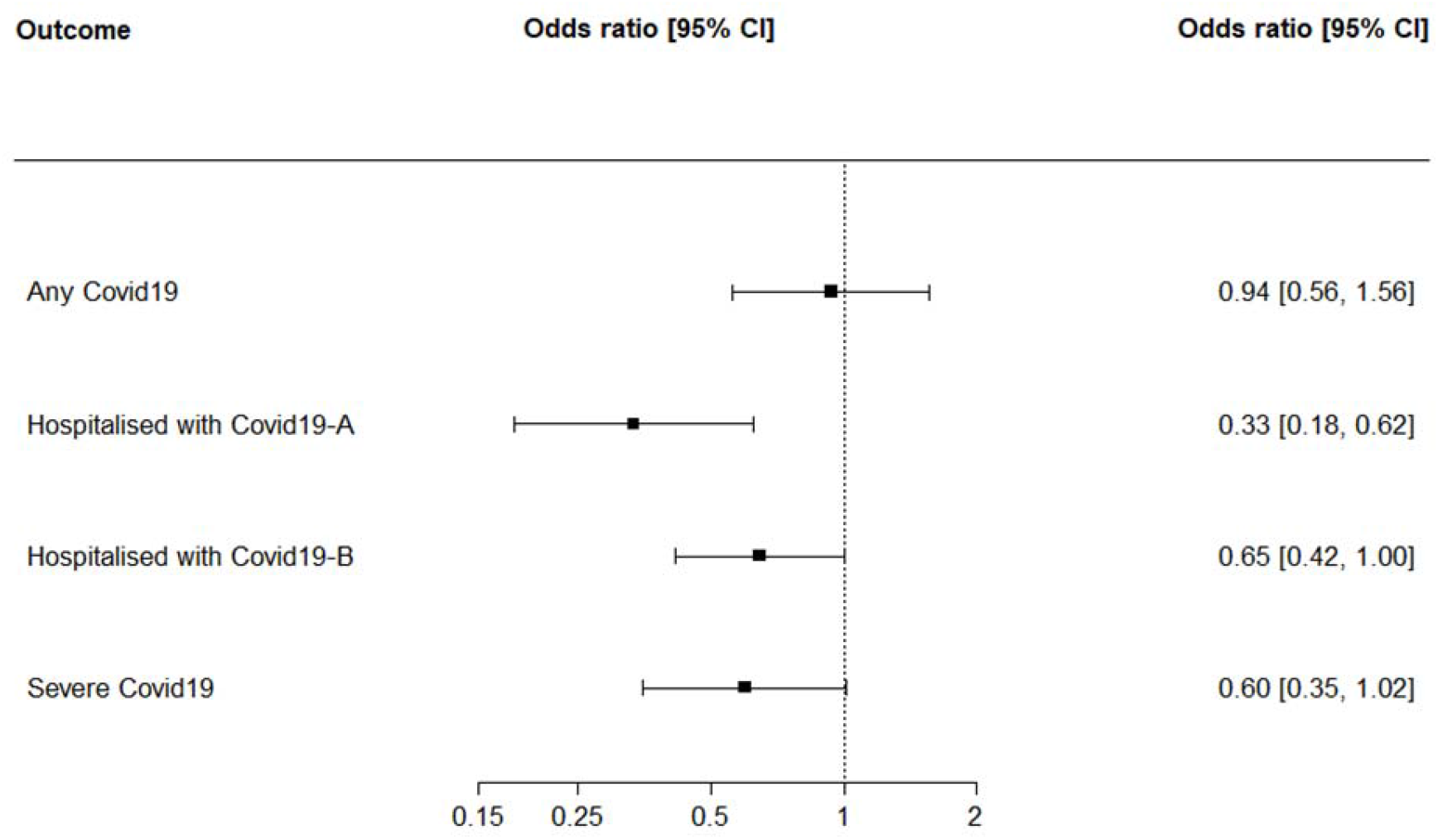
Forest plot showing odds ratio estimates from Mendelian Randomization analyses of acetyl-carnitine concentration against Covid19 outcomes. Hospitalised with Covid19-A = hospitalised with Covid19 versus infected but not hospitalised with Covid 19; Hospitalised with Covid19-B = hospitalised with Covid19 versus population controls.

### Carnitine and Covid19

Our MR results for the effect of carnitine on severe Covid19 were consistent with the acetyl-carnitine results (Table 1; Figure 2 and supplemental material; Figures S2-S3), but there was stronger evidence of association with any Covid19 infection and weaker evidence for an effect on hospitalisation with Covid19 in our main analysis. We found evidence that a doubling in blood carnitine levels reduces the risk of any Covid19 infection by 19%, being hospitalised with Covid19 by 18% (versus infected but not hospitalised) and 20% (compared to population controls) and reduces risk of severe Covid19 by 44%. However, in all sensitivity analyses (supplementary table S5) results showed much stronger evidence and greater protective effects of carnitine on hospitalisation with Covid19 than our main analyses.

**Figure 2.**
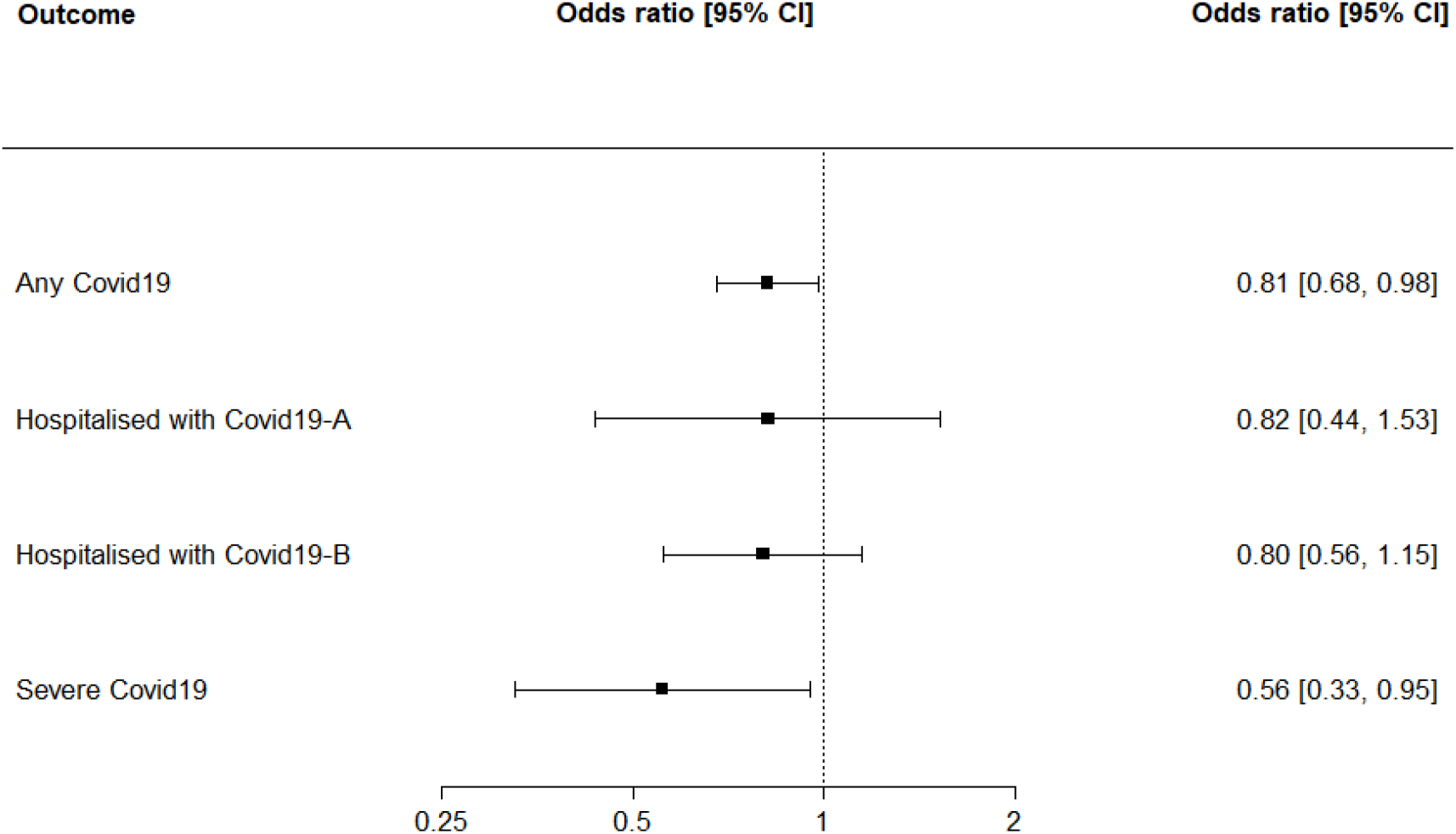
Forest plot showing odds ratio estimates from Mendelian Randomization analyses of carnitine concentration against Covid19 outcomes. Hospitalised with Covid19-A = hospitalised with Covid19 versus infected but not hospitalised with Covid 19; Hospitalised with Covid19-B = hospitalised with Covid19 versus population controls.

There was no strong evidence of heterogeneity between SNPs for carnitine and any of the Covid19 outcomes. Among the outcomes found to be associated with our carnitine SNPs were all those associated with the acetyl-carnitine SNPs, plus the following outcomes which were associated with at least 1 SNP at GWAS significance level; age first had intercourse, testosterone, lipids, betaine, C-reactive protein, phosphate, selenium, cystatin C, alanine, forced expiratory volume, mean arterial pressure, hearing difficulty and heel bone mineral density (supplemental material; Table S2). Leave one out analyses indicated that our results were not driven by a single SNP. Steiger test showed no evidence that our secondary outcomes affected carnitine or acetyl-carnitine levels in these analyses.

### Secondary outcomes

To determine the impact of acetyl-carnitine and carnitine on our positive control outcome (muscle weakness) and other comorbidities, we performed MR analyses of acetyl-carnitine and carnitine on a range of health outcomes (Supplemental material; Table S6-S9 and Figure S4-S5). We had statistical power of ≥75% to detect OR of 1.2 (or 0.80) (Supplemental material; Table S3) per doubling of exposure in these analyses.

The results showed evidence of positive association between acetyl-carnitine and BMI with a tenth of a BMI unit increase per doubling of acetyl-carnitine (beta; 0.05 95% CI: 0.00 to 0.11 and P=0.05) (supplemental material; Table S7), we found a weak inverse effect of acetylcarnitine on fat mass and an inverse effect on fat-free mass. There was also an inverse relationship between acetylcarnitine and height. Only one acetyl-carnitine SNP was available in the GWAS of C-reactive protein, which showed an inverse relationship with C-reactive protein levels (beta_natural log unit change per doubling of acetylcarnitine_; -0.18 95%CI -0.30 to -0.06 p=0.003). There was very strong evidence of heterogeneity between the two SNPs for height and fat free mass, although the effects for both were in the same direction. For the remaining secondary outcomes the evidence of association was not strong. The results showed modest evidence of positive association with coronary heart disease (OR: 1.36; 95% CI: 0.97 to 1.92 and P=0.08). We found heterogeneity between SNPs (P_het_ ≤ 0.05) for breast cancer and muscle weakness but due to small number of SNPs we were not able to perform sensitivity analyses (Supplemental material; Table S6, Figure S4).

For carnitine our IVW analysis we found evidence of positive association with coronary heart disease (OR: 1.43; 95% CI: 1.07 to 1.90 and P=0.01) and type 2 diabetes (OR: 1.35; 95% CI: 1.04 to 1.75 and P=0.02), although the coronary heart disease finding attenuated in sensitivity analyses (Supplemental material; Table S8 and Figure S5). The outcomes BMI, breast cancer, prostate cancer, T2D, muscle weakness and stroke showed evidence of heterogeneity between SNPs. MR-PRESSO analyses were executed for these outcomes to identify the outlying SNPs and the results detected outliers for outcomes except prostate cancer. After removing the outlying SNPs, the analyses were re-ran and the IVW analyses are presented (Supplemental material; Figure S4 and Table S8). The results of our analysis of carnitine and fat mass, fat-free mass and height reflected those for acetylcarnitine (supplemental material; Table S9), with inverse associations for all these outcomes, but with the largest effect on height. We also observed a great deal of heterogeneity between the SNPs for anthropometry measures (especially height) with several SNPs showing large effects in each direction but removal of outliers did not substantially change our effect estimates. We did not find strong evidence of an association of carnitine with C-reactive protein levels overall, although again there was substantial heterogeneity in this analysis.

Effect estimates in our multivariable Mendelian randomization (MVMR) analyses did not differ substantially from our main analyses (supplemental material; Table S9), the MVMR analysis of acetyl-carnitine and diabetes on severe Covid19 was attenuated compared to the main analysis but this was because only one SNP was included in the MVMR analysis whereas there were two SNPs in the main analysis.

## Discussion

Using common genetic polymorphisms as proxies for carnitine and acetyl-carnitine exposure we found evidence that both metabolites protected against severe Covid19 infection. We found around a 40% reduction of this outcome associated with a doubling of either acetyl-carnitine or carnitine. For carnitine analyses where hospitalisation or severe Covid19 were the outcomes our sensitivity analyses showed stronger protective effects than our IVW analyses, this was particularly true for MR-Egger which tests whether the SNP-outcome effect is proportionate to the SNP-exposure effect (dose response). Thus our IVW analyses where hospitalisation with Covid19 is the outcome may be influenced by pleiotropy (effects of genetic variants on Covid19 which are independent of carnitine) but this is more likely to be masking an effect of carnitine rather than inflating it. Although our MVMR analyses with adjustment for BMI, diabetes and heart disease, did not substantially change our estimates.

We did not find robust effects of carnitine or acetyl-carnitine on Covid19 infection (*any Covid19*), suggesting that these molecules are not having an impact on susceptibility to infection but on disease severity. However, because population controls were used, our analyses of the effect of carnitine on susceptibility to infection may be biased if when those with high carnitine levels become infected they are less likely to show symptoms and receive a Covid19 diagnosis.

For acetyl-carnitine our estimates suggested stronger protective effects for hospitalisation with Covid19 when comparing with those who had Covid19 infection but were not hospitalised, this is consistent with our expectation because the majority of people in the population control group will not have been infected with Covid19 including some individuals who may be hospitalised or have severe Covid19 if infected. We were unable to run sensitivity analyses for acetyl-carnitine due to the small number of genetic instruments (n=2), but the instruments gave consistent estimates in analyses where severe Covid19 or hospitalization from Covid19 were the outcomes. To investigate the potential for pleiotropy we explored other outcomes associated with these SNPs. The outcomes we identified, anthropometry, other carnitines and inflammatory diseases, were likely to be downstream effects of acetyl-carnitine and some such as height must have been influenced by carnitine early in life. We found some heterogeneity in the magnitude of effects (even accounting for differential effects on acetylcarnitine) but consistency in the traits associated with the two SNPs.

### Consistency with other evidence

Carnitine has been shown to be effective at counteracting proinflammatory conditions and reducing oxidative stress in animal models and has therefore been hypothesized to be a potentially effective treatment for Covid19 induced pneumonia and sepsis.^35^ In a meta-analysis of randomized controlled trials which examined the effects of supplementation with L-carnitine on oxidative stress and inflammatory proteins carnitine supplementation resulted in a lowering of CRP, IL-6 and TNF-α.^36^ In our analysis we did not find strong evidence of an effect of our carnitine instrument on CRP, although there was substantial heterogeneity in this analysis. We did find that acetyl-carnitine reduced CRP levels in blood but only one acetylcarnitine SNP was represented in CRP GWAS. A recent preprint describes a randomized controlled trial in which 304 patients with mild to moderate Covid19 were randomized to receive combined metabolic activators including carnitine.^37^ The trial found that the time it took to become symptom free was 6.6 days in those receiving metabolic activators compared with 9.3 days in those receiving placebo.^38^

### Positive controls and comorbidities

We found heterogeneous effects in our MR analyses of carnitine and acetyl-carnitine SNPs with muscle weakness, which was unexpected given that carnitine deficiency at birth is associated with muscle weakness. A possible explanation is that the outcome used in our analysis was measured in elderly adults not children. An alternative explanation is that we were proxying blood carnitine/acetyl-carnitine levels in our analyses and the SNPs we used may have more heterogeneous effects on carnitine levels in muscle tissue. The latter hypothesis is supported by our findings that the acetyl-carnitine increasing allele in SNP rs274567 which maps to the carnitine transporter gene *SLC22A5(OCTN2)* increased risk of muscle weakness by around 70% (OR=1.68, 95%CI 1.30 to 2.16 p=0.00007; Supplemental Material Figure S4) and decreased handgrip strength in the UKBiobank study (Supplemental material; Table S4). This finding is consistent with inhibition of carnitine uptake in tissues by high levels of acetyl-carnitine in blood, which has been previously described^38,39^ and could be part of a negative feedback loop arising from a reduced uptake capacity of *SLC22A5* associated with rs274567.

We found some evidence that both carnitine and acetyl-carnitine increased the risk of coronary heart disease and type 2 diabetes. However our result for coronary heart disease was attenuated in sensitivity analyses. Several randomized controlled trials have investigated whether carnitine is an effective treatment to prevent further morbidity and mortality among those who have had a myocardial infarction. A systematic review and meta-analysis of such trials found that L-carnitine reduced the risk of angina and ventricular arrhythmias, but did not find strong evidence of a protective effect on heart failure or reinfarction.^40^ This systematic review has been heavily criticised for its methodology and the inclusion of a previously discredited study.^41^ Other studies have found that supplementation with L-carnitine leads to an increase in trimethylamine-N-oxide (TMAO) which can lead to atherosclerosis.^42,43^ In agreement with this finding two large cohort studies of high risk individuals have shown that TMAO levels at baseline positively associated with a major acute coronary event during follow-up.^44^ Two separate cohort studies of high risk individuals found that higher levels of acylcarnitines at baseline were associated with a higher risk of cardiovascular death at follow-up.^45 46^

Inborne errors of metabolism which lead to carnitine deficiencies lead to hypoglycaemia and low levels of ketones in blood due to an inability to metabolise fatty acids for energy.^23^ High levels of carnitine might therefore be expected to lead to hyperglycaemia and an increased risk of type 2 diabetes as was seen in our analysis. In agreement with our findings several cohort studies have found carnitine and acylcarnitines to be associated with an increase in type 2 diabetes risk.^47-49^Although other studies have found that carnitine supplementation can have beneficial effects on lipid profiles among type 2 diabetic patients.^50^

We found a predicted causal effect of acetyl-carnitine on increased BMI whereas we did not find such an effect for carnitine. However, both carnitine and acetyl-carnitine showed inverse relationships with fat-free mass. Stronger inverse relationships were apparent with both carnitine and acetyl-carnitine and height, suggesting that the effect of acetyl-carnitine and BMI may reflect growth in childhood, since height is fixed in adults. A recent systematic review and meta-analysis of randomized controlled trials found that carnitine supplementation could help weight loss,^51^ although results were heterogeneous between the trials and studies were not assessed for quality which is an important omission given that several trials did not include a placebo and there was evidence of publication bias for the BMI result. We didn’t find any trials of acetyl-carnitine which measured body weight as a outcome, but we did find a small observational study which reported a positive association between fat mass and plasma acetyl-carnitine levels.^52^ Secondary carnitine deficiency can arise in people with kidney failure and other metabolic disorders,^23^ so we tested the direction of association using in our analyses of kidney disease, coronary heart disease, type II diabetes and anthropometry. In all analyses there was strong evidence that the true direction of effect was from carnitine/acetyl-carnintine to outcome.

In summary, acetyl-carnitine and carnitine may increase coronary heart disease and type II diabetes which are associated with severe Covid19, and acetyl-carnitine appears to increase BMI (although this is likely due to early effects on growth). If anything these off target effects are likely to reduce the magnitude of the protective effect of carnitine and acetylcarnitine on Covid19 severity in our analysis, although we did not find evidence for this in our analyses. However, they are an important consideration when decided whether carnitines are a safe treatment for those with Covid19. Carnitine supplementation is widely viewed as safe, it is available as a supplement from health food shops, used by atheletes,^52-54^ those on hemodialysis^55^ and cancer patients.^56^ However trials investigating their safety have tended to be too small to detect rare effects.^524-56^

### Strengths and limitations of our analyses

Randomized controlled trials are expensive and are unethical where evidence of the treatment effect is weak, particularly if the drug in question has potential off target effects. In addition, trials take time and in the context of a global pandemic of a novel virus prioritization of the most promising treatments to take forward into trials is essential. Two sample Mendelian randomization analyses, such as the ones we have performed, can make use of large-scale data on common genetic variants from existing genome wide association studies,^57^ which means the analyses can be carried out quickly and can inform trials. In addition, using Mendelian randomization it is possible to look at effects of the treatment or exposure of interest with other outcomes using data from different GWAS to investigate whether they are on the causal pathway and to estimate any potential off-target effects.

An important interpretive issue with Mendelian randomization is that genetic variants generally reflect variation in exposure over a lifetime so it is not possible to determine with certainty from our analyses the effect of short-term treatment with acetyl-carnitine and carnitine for the prevention of severe Covid19. Some of the effects we found with our secondary outcomes clearly occurred early in life (height) and are therefore very unlikely to be influenced by treatment of adults with carnitine. In addition, our analysis assumed a linear effect of the exposure over all levels and does not provide information on whether there is a threshold effect; it is possible that only individuals who are deficient in carnitine would benefit from treatment.

In summary, we found strong protective effects of both blood acetyl-carnitine and carnitine levels on severe Covid19. Carnitine and acetyl-carnitine may therefore be effective treatments in those who have contracted Covid19. However, we also find that genetic predisposition to higher carnitines may increase both coronary heart disease and type 2 diabetes risk and BMI. It is plausible that short term treatment with carnitines will have beneficial effects of reducing inflammation and severe Covid19 without causing long term harm and it is worth formally evaluating whether the balance of evidence would support incorporating carnitine in RCTs of Covid-19 treatment.

## Data Availability

All the data included in this manuscript is publicly available.

## Acknowledgements

We would like to thank the Covid19 GWAS consortium and the authors of all the GWASs who made their summary statistics available for the benefit of this and similar work. We would also like to thank our colleagues responsible for setting up the MRBase platform, which has made it possible to carry-out this research in such a short time period. Dr Kazmi would like to thank her husband Imran Kazmi for his constant encouragement and support throughout this research.

