## Supplemental material for "Mendelian randomization analyses show that higher acetyl-carnitine and carnitine levels in blood protect against severe Covid19"

**Table S1: The GWAS summary statistics for the effect of two acetyl-carnitine SNPs on blood acetyl-carnitine levels, taken from Shin et al.^1^**

| **SNP** | **CHR** | **Gene** | **Position** | **EA** | **OA** | **EAF** | **Beta** | **SE** | **P** | **Other outcomes** |
| --- | --- | --- | --- | --- | --- | --- | --- | --- | --- | --- |
| rs274567 | 5 | SLC22A5 | 131714409 | T | C | 0.37 | 0.018 | 0.0025 | 2.85E-12 | Eosinophil counts (PMID: 27863252), height (PMID: 25282103), appendicular lean mass (PMID: 33097823), body mass (UK Biobank), diastolic blood pressure (PMID: 30224653),  Crohn’s disease (PMID: 26192919), propionyl-carnitine (PMID: 24816252), hand grip strength (UK Biobank), oleoyl-carnitine (PMID: 24816252), hexanoyl-carnitine (PMID: 24816252), inflammatory bowel disease (PMID: 26192919), asthma (UK Biobank), neutrophil count (PMID: 27863252) |
| rs1171614 | 10 | SLC16A9 | 61469538 | C | T | 0.78 | 0.028 | 0.0028 | 3.38E-23 | Height (UK Biobank), propionylcarnitine (PMID: 24816252, Urate (PMID: 23263486), butyrylcarnitine (PMID: 24816252), appendicular lean mass (PMID: 33097823),  self-reported gout (UK Biobank), body mass (UK Biobank), hexanoylcarnitine (PMID: 24816252), triglycerides (UK biobank), treatment/medication code: allopurinol (UK Biobank), basal metabolic rate (UK Biobank) |

The GWAS summary statistics for the effect of two acetyl-carnitine SNPs on blood acetyl-carnitine levels, taken from Shin et al.^1^ EA = the allele that is associated with higher acetyl-carnitine levels, OA = other allele, EAF = effect allele frequency, Beta= the beta coefficient for the effect on acetyl-carnitine levels on a log10 scale, SE = standard error for effect of the SNP on acetyl-carnitine levels on a log10 scale, P= p-value for effect on acetylcarnitine levels, Other outcomes = other traits associated with carnitine SNPs at p<5×10^-8^

| **SNP** | **CHR** | **Gene** | **Position** | **EA** | **OA** | **EAF** | **Beta** | **SE** | **P** | **Associated outcomes** |
| --- | --- | --- | --- | --- | --- | --- | --- | --- | --- | --- |
| rs1466788 | 1 | Unknown | 110618730 | G | A | 0.59 | 0.007 | 0.0009 | 3.05E-16 | Age first had sexual intercourse (UK Biobank) |
| rs735315 | 2 | LOC730100 | 51331157 | T | C | 0.49 | 0.005 | 0.0009 | 1.87E-09 | None |
| rs2279014 | 2 | SLC11A1 | 219261176 | C | T | 0.63 | 0.005 | 0.0009 | 4.86E-08 | Height (PMID: 25282103), platelet count/distribution (UK Biobank), erythrocyte distribution (UK Biobank), |
| rs9842133 | 3 | PEX5L | 179664102 | T | C | 0.67 | 0.006 | 0.0009 | 4.20E-12 | None |
| rs4860022 | 4 | SPOCK3 | 167693875 | C | T | 0.66 | 0.006 | 0.0009 | 9.60E-10 | None |
| rs419291 | 5 | SLC22A4 | 131633355 | T | C | 0.37 | 0.008 | 0.0009 | 3.10E-18 | Blood cell count (Blood Cell Consortium), height (PMID: 25282103),  appendicular lean mass (PMID: 33097823), isovalerylcarnitine (PMID: 24816252), body mass (UK Biobank),  basal metabolic rate (UK Biobank), Crohn’s disease (PMID: 26192919), diastolic blood pressure (PMID: 30224653),  hand grip strength (UK Biobank), inflammatory bowel disease (PMID: 26192919), asthma (UK Biobank), FEV (UK Biobank) |
| rs13182512 | 5 | JMY | 78573790 | T | A | 0.62 | 0.005 | 0.0009 | 3.36E-08 | Triglycerides (PMID: 32203549), selenium (PMID: 23720494), HDL cholesterol (PMID: 32203549), betaine (PMID: 24816252), CRP (UK Biobank) |
| rs6862024 | 5 | TNIP1 | 150428871 | G | A | 0.63 | 0.006 | 0.0009 | 8.99E-10 | None |
| rs2396004 | 6 | Unknown | 43355851 | A | G | 0.44 | 0.005 | 0.0009 | 4.31E-08 | Cystatin C (UK Biobank), Urate (UK Biobank), systolic blood pressure (PMID: 30224653), hearing difficulty (UK Biobank), high blood pressure (UK Biobank),  testosterone (UK Biobank), platelet crit (UK Biobank), height (PMID: 26367797), uric acid (PMID: 29403010), mean arterial pressure (PMID: 29403010),  heel bone mineral density (PMID: 30598549) |
| rs10821585 | 10 | Unknown | 61516587 | G | A | 0.59 | 0.009 | 0.0009 | 1.28E-21 | Urate (UK Biobank), gout (UK Biobank) |
| rs12356193 | 10 | SLC16A9 | 61413353 | A | G | 0.84 | 0.027 | 0.0016 | 3.69E-63 | Urate (UK Biobank), height (UK Biobank), triglycerides (PMID: 32203549), testosterone (UK Biobank), gout (UK Biobank) |
| rs11183620 | 12 | SLC38A4 | 47212370 | A | G | 0.47 | 0.005 | 0.0009 | 3.00E-08 | Height (UK Biobank), alanine (PMID: 27005778) |
| rs11620955 | 14 | FNTB/MAX/CHURC1-FNTB | 65491255 | G | A | 0.52 | 0.005 | 0.0009 | 1.63E-08 | Mean corpuscular haemoglobin (PMID: 27863252), height (UK Biobank), reticulocyte percentage (UK Biobank), platelet count (UK Biobank), body mass (UK Biobank),  hand grip strength (UK Biobank) |
| rs11620973 | 14 | Unknown | 96018052 | G | A | 0.37 | 0.005 | 0.0009 | 2.35E-08 | None |
| rs2114713 | 15 | CTXND1 | 80528373 | G | T | 0.42 | 0.005 | 0.0009 | 2.67E-08 | None |
| rs3736438 | 17 | EFCAB13 | 45486651 | G | T | 0.61 | 0.006 | 0.0009 | 3.19E-10 | Low density lipoprotein cholesterol levels (PMID: 32493714), apolipoprotein B (PMID: 32203549), LDL cholesterol (PMID: 32203549),  height (UK Biobank), cholesterol (UK Biobank), phosphate (UK Biobank), urate (UK Biobank) |
| rs12709393 | 17 | Unknown | 55292115 | T | G | 0.63 | 0.005 | 0.0009 | 6.41E-09 | None |

**Table S2: The GWAS summary statistics for the effect of seventeen carnitine SNPs on blood carnitine levels, taken from Shin et al.**

**^1^**The GWAS summary statistics for the effect of seventeen carnitine SNPs on blood carnitine levels, taken from Shin et al.^1^EA **=** the allele that is associated with higher carnitine levels, OA = other allele, EAF = effect allele frequency, Beta= the beta coefficient for the effect on carnitine levels on a log10 scale, SE = standard error for effect of the SNP on carnitine levels on a log10 scale, P= p-value for effect on carnitine levels, associated outcomes = other traits associated with carnitine SNPs at p<5×10^-8^

**Table S3. Details of the GWAS from which summary statistics for our secondary outcomes were taken.**

| **Trait** | **PMID/MR-Base id** | **Consortium/Dataset** | **Sample size** | **Cases** | **Sex** | **Unit** | **^a^Power (%)** | **^b^Power (%)** |
| --- | --- | --- | --- | --- | --- | --- | --- | --- |
| Alzheimer’s disease^2^ | 30617256 | ADSP, PGC-ALZ and UKB | 455258 | 71880 | Males and females | Z- unit change | >99 | >99 |
| Body Mass Index^3^ | 30124842  Ukb-b-2303* | GIANT (a)  UKBiobank (b) | 681275  454884 | - | Males and females | kg/m^2^ | >99 | >99 |
| Breast cancer^4^ | 29059683 | BCAC | 228951 | 122977 | Females | Log odds ratio | >99 | >99 |
| Coronary heart Disease^5^ | 21378990 | CARDIoGRAM | 86995 | 22233 | Males and females | Log odds ratio | 90 | >99 |
| Chronic kidney disease^6^ | 26831199 | GWAS meta-analysis included participants from 43 studies | 117165 | 12385 | Males and females | Log odds ratio | 75 | >99 |
| C-reactive protein^7^ | 30388399 | GWAS meta-analysis included participants from 88 studies | 204402 | - | Males and females | Natural log (mg/L) | >99 | >99 |
| Height^8^ | 25282103 | GIANT | 253288 | - | Males and females | SD (m) | >99 | >99 |
| Muscle weakness^9^ | 33510174 | GWAS meta-analysis included participants from 22 studies | 256523 | 48596 | Males and females | Grip strength <30 kg Male; <20 kg Female | >99 | >99 |
| Prostate cancer^10^ | 29892016 | PRACTICAL | 140254 | 79148 | Males | Odds ratio | >77 | >99 |
| Stroke^11^ | 29531354 | GWAS meta-analysis included participants from 17 studies | 446696 | 40585 | Males and females | Log odds ratio | >99 | >99 |
| Type II Diabetes^12^ | 30054458 | DIAGRAM, GERA and UKB | 655666 | 61714 | Males and females | Log odds ratio | >93 | >99 |
| *Whole body fat free mass | ukb-b-1335* | UK Biobank | 454850 | - | Males and females | SD (kg) | >99 | >99 |
| *Whole body fat mass | ukb-b-19393* | UK Biobank | 454137 | - | Males and females | SD (kg) | >99 | >99 |

Details of the GWAS from which summary statistics for our secondary outcomes were taken. ^a^Power = power to detect an odds ratio of 1.2(0.8) for an association of the acetyl-carnitine with the outcome at a significance level (P) of 0.05; ^b^Power = power to detect an odds ratio of 1.2(0.8) for an association of the carnitine with the outcome at a significance level (P) of 0.05.

*MR-Base ids are provided for these outcomes.

Two different GWAS were used for the BMI analyses, the GIANT consortium (a) was used for the carnitine analysis as this was the largest available GWAS and included all 17 carnitine SNPs, however UKBiobank (b) was used for acetylcarnitine as this was the largest available GWAS which included both acetylcarnitine SNPs.

**Table S4. Genetic associations of acetyl-carnitine SNPs for other outcomes identified as being associated with at least one SNP in our instrument in genome wide association studies**

| **SNP** | **Outcome** | **PMID/MR Base id** | **Beta** | **Se** | **P** | **Sample size** |
| --- | --- | --- | --- | --- | --- | --- |
| rs274567 | *Crohn's disease | 26192919 | -0.087 | 0.012 | 3.25E-12 | 51874 |
| rs274567 | *Diastolic blood pressure | 30224653 | 0.125 | 0.018 | 3.16E-12 | 747899 |
| rs274567 | *Eosinophil cell count | **ieu-b-33 | 0.050 | 0.002 | 5.04E-131 | 471755 |
| rs274567 | *Inflammatory bowel disease | 26192919 | -0.065 | 0.010 | 2.13E-10 | 65642 |
| rs274567 | *Monocyte cell count | **ieu-b-31 | 0.021 | 0.002 | 3.74E-28 | 519114 |
| rs274567 | *Neutrophil cell count | **ieu-b-34 | 0.016 | 0.002 | 1.16E-15 | 516806 |
| rs274567 | *White blood cell count | **ieu-b-30 | 0.026 | 0.002 | 2.04E-42 | 556269 |
| rs274567 | 2-tetradecenoyl carnitine | 24816252 | 0.026 | 0.004 | 1.27E-09 | 6595 |
| rs1171614 | 2-tetradecenoyl carnitine | 24816252 | 0.008 | 0.006 | 0.16 | 6595 |
| rs274567 | Allopurinol | **ukb-b-15895 | -0.0002 | 0.0002 | 0.36 | 462933 |
| rs1171614 | Allopurinol | **ukb-b-15895 | 0.002 | 0.0003 | 9.10E-11 | 462933 |
| rs274567 | Appendicular lean mass | 33097823 | -0.023 | 0.002 | 6.74E-32 | 450243 |
| rs1171614 | Appendicular lean mass | 33097823 | -0.017 | 0.002 | 1.13E-13 | 450243 |
| rs274567 | Asthma | **ukb-a-446 | 0.005 | 0.001 | 1.38E-09 | 336782 |
| rs1171614 | Asthma | **ukb-a-446 | 0.0009 | 0.001 | 0.83 | 336782 |
| rs274567 | Basal metabolic rate | **ukb-b-16446 | -0.010 | 0.001 | 2.50E-14 | 454874 |
| rs1171614 | Basal metabolic rate | **ukb-b-16446 | -0.009 | 0.002 | 4.10E-08 | 454874 |
| rs274567 | Butyryl-carnitine | 24816252 | 0.024 | 0.004 | 6.23E-09 | 7349 |
| rs1171614 | Butyryl-carnitine | 24816252 | 0.035 | 0.005 | 6.78E-14 | 7349 |
| rs274567 | Granulocyte count | 27863252 | 0.025 | 0.004 | 1.11E-11 | 169822 |
| rs1171614 | Granulocyte count | 27863252 | 0.006 | 0.004 | 0.19 | 169822 |
| rs274567 | Hand grip strength (right) | **ukb-b-10215 | -0.010 | 0.002 | 4.00E-11 | 461089 |
| rs1171614 | Hand grip strength (right) | **ukb-b-10215 | -0.002 | 0.002 | 0.23 | 461089 |
| rs274567 | Height | 25282103 | -0.034 | 0.003 | 4.20E-30 | 253200 |
| rs1171614 | Height | 25282103 | -0.021 | 0.004 | 1.20E-07 | 231866 |
| rs274567 | Hexanoyl-carnitine | 24816252 | 0.022 | 0.003 | 5.86E-11 | 7340 |
| rs1171614 | Hexanoyl-carnitine | 24816252 | 0.025 | 0.004 | 4.05E-11 | 7340 |
| rs274567 | Isovaleryl-carnitine | 24816252 | 0.038 | 0.003 | 4.64E-31 | 7344 |
| rs1171614 | Isovaleryl-carnitine | 24816252 | 0.009 | 0.004 | 0.02 | 7344 |
| rs274567 | Mean platelet volume | 27863252 | -0.036 | 0.004 | 9.97E-22 | 164454 |
| rs1171614 | Mean platelet volume | 27863252 | -0.012 | 0.004 | 0.005 | 164454 |
| rs274567 | Oleoyl-carnitine | 24816252 | 0.023 | 0.004 | 5.65E-11 | 7263 |
| rs1171614 | Oleoyl-carnitine | 24816252 | 0.012 | 0.005 | 0.01 | 7263 |
| rs274567 | Palmitoyl-carnitine | 24816252 | 0.021 | 0.003 | 5.88E-10 | 7259 |
| rs1171614 | Palmitoyl-carnitine | 24816252 | 0.008 | 0.004 | 0.04 | 7259 |
| rs274567 | Propionyl-carnitine | 24816252 | 0.017 | 0.003 | 3.47E-11 | 7364 |
| rs1171614 | Propionyl-carnitine | 24816252 | 0.032 | 0.003 | 2.49E-29 | 7364 |
| rs274567 | Self-reported gout | **ukb-b-13251 | -0.0005 | 0.0003 | 0.05 | 462933 |
| rs1171614 | Self-reported gout | **ukb-b-13251 | 0.002 | 0.0003 | 3.60E-17 | 462933 |
| rs274567 | Treatment/medication code: allopurinol | **ukb-b-15895 | -0.0002 | 0.0002 | 0.36 | 462933 |
| rs1171614 | Treatment/medication code: allopurinol | **ukb-b-15895 | 0.002 | 0.0003 | 9.10E-11 | 462933 |
| rs274567 | Triglycerides | **ukb-d-30870_irnt | 0.009 | 0.002 | 0.0001 | 343992 |
| rs1171614 | Triglycerides | **ukb-d-30870_irnt | 0.018 | 0.003 | 3.20E-11 | 343992 |
| rs274567 | Urate | 23263486 | -0.012 | 0.006 | 0.05 | 110037 |
| rs1171614 | Urate | 23263486 | 0.074 | 0.007 | 6.48E-23 | 103697 |
| rs274567 | Whole body fat-free mass | **ukb-b-13354 | -0.010 | 0.001 | 9.50E-16 | 454850 |
| rs1171614 | Whole body fat-free mass | **ukb-b-13354 | -0.009 | 0.001 | 2.30E-09 | 454850 |
| rs274567 | Whole body water mass | **ukb-b-14540 | -0.01 | 0.001 | 3.60E-16 | 454888 |
| rs1171614 | Whole body water mass | **ukb-b-14540 | -0.009 | 0.001 | 2.60E-09 | 454888 |

*Only one of the two SNPs of acetyl-carnitine concentration was available for these outcomes.

**PMIDs were not available therefore MR-Base ids are provided.

**Table S5. Result of our main and sensitivity MR analyses of carnitine on Covid19 outcomes.**

| **Outcome** | **Method** | **# SNPs** | **OR** | **LCI** | **UCI** | **P** |
| --- | --- | --- | --- | --- | --- | --- |
| Any Covid19 | Inverse variance weighted | 17 | 0.81 | 0.68 | 0.98 | 0.03 |
| Any Covid19 | MR Egger | 17 | 0.93 | 0.66 | 1.32 | 0.70 |
| Any Covid19 | Weighted median | 17 | 0.85 | 0.67 | 1.08 | 0.19 |
| Any Covid19 | Weighted mode | 17 | 0.86 | 0.66 | 1.11 | 0.27 |
| Hospitalised with Covid19 (A) | Inverse variance weighted | 17 | 0.82 | 0.44 | 1.53 | 0.52 |
| Hospitalised with Covid19 (A) | MR Egger | 17 | 0.26 | 0.09 | 0.79 | 0.03 |
| Hospitalised with Covid19 (A) | Weighted median | 17 | 0.46 | 0.20 | 1.02 | 0.06 |
| Hospitalised with Covid19 (A) | Weighted mode | 17 | 0.40 | 0.16 | 0.98 | 0.06 |
| Hospitalised with Covid19 (B) | Inverse variance weighted | 17 | 0.80 | 0.56 | 1.15 | 0.23 |
| Hospitalised with Covid19 (B) | MR Egger | 17 | 0.48 | 0.25 | 0.93 | 0.05 |
| Hospitalised with Covid19 (B) | Weighted median | 17 | 0.63 | 0.39 | 1.02 | 0.06 |
| Hospitalised with Covid19 (B) | Weighted mode | 17 | 0.60 | 0.36 | 1.01 | 0.07 |
| Severe Covid19 | Inverse variance weighted | 17 | 0.56 | 0.33 | 0.95 | 0.03 |
| Severe Covid19 | MR Egger | 17 | 0.41 | 0.16 | 1.07 | 0.09 |
| Severe Covid19 | Weighted median | 17 | 0.50 | 0.25 | 1.00 | 0.05 |
| Severe Covid19 | Weighted mode | 17 | 0.51 | 0.26 | 1.00 | 0.07 |

Hospitalised with Covid19-A = Hospitalised Covid19 vs. not hospitalised Covid19; Hospitalised with Covid19-B =. Hospitalised Covid19 vs. population

**Table S6. MR results of acetyl-carnitine on Covid19 related comorbidities (binary outcomes).**

| **Outcome** | **# SNPs** | **Method** | **OR** | **LCI** | **UCI** | **P_assoc_** | **P_het_** |
| --- | --- | --- | --- | --- | --- | --- | --- |
| Alzheimer’s | 2 | Inverse variance weighted | 1.00 | 0.93 | 1.06 | 0.89 | 0.15 |
| *Breast cancer | 2 | Inverse variance weighted | 0.93 | 0.57 | 1.52 | 0.78 | 0.0002 |
| Breast cancer-rs1171614 | 1 | Wald ratio | 1.13 | 0.96 | 1.34 | 0.14 | - |
| Breast cancer-rs274567 | 1 | Wald ratio | 0.68 | 0.55 | 0.84 | 3.9×10^-4^ | - |
| Coronary heart disease | 2 | Inverse variance weighted | 1.36 | 0.97 | 1.92 | 0.08 | 0.46 |
| Chronic kidney disease | 2 | Inverse variance weighted | 1.05 | 0.64 | 1.72 | 0.86 | 0.10 |
| *Muscle weakness | 2 | Inverse variance weighted | 1.16 | 0.69 | 1.96 | 0.58 | 0.0005 |
| Muscle weakness-rs1171614 | 1 | Wald ratio | 0.95 | 0.79 | 1.15 | 0.62 | - |
| Muscle weakness-rs274567 | 1 | Wald ratio | 1.68 | 1.30 | 2.16 | 7.08×10^-5^ | - |
| Prostate cancer | 1 | Wald ratio | 0.81 | 0.62 | 1.06 | 0.13 | - |
| Stroke | 2 | Inverse variance weighted | 0.97 | 0.80 | 1.17 | 0.72 | 0.57 |
| Type II diabetes | 1 | Wald ratio | 1.18 | 0.90 | 1.55 | 0.24 | - |

OR = odds ratio; LCI = lower 95% confidence interval; UCI = upper 95% confidence interval; P_assoc_ = P-value for association; P_het_ = P-value for heterogeneity between instrumental SNP causal estimates; # SNPs = total number of SNPs used in the analysis

*The outcomes showed evidence of heterogeneity between SNPs (Phet ≤ 0.05) therefore results of single SNP MR analyses are also presented.

**Table S7. MR results of acetyl-carnitine on Covid19 related comorbidities (continuous outcomes).**

| **Outcome** | **Method** | **# SNPs** | **Beta** | **LCI** | **UCI** | **P_assoc_** | **P_het_** |
| --- | --- | --- | --- | --- | --- | --- | --- |
| Body Mass Index | Inverse variance weighted | 2 | 0.05 | 0.00 | 0.11 | 0.05 | 0.19 |
| C-reactive protein | Wald ratio | rs274567 | -0.18 | -0.30 | -0.06 | 0.003 | - |
| *Height | Inverse variance weighted | 2 | -0.37 | -0.71 | -0.03 | 0.03 | 8.85×10^-8^ |
| Height | Wald ratio | rs1171614 | -0.23 | -0.31 | -0.14 | 7.26×10^-8^ | - |
| Height | Wald ratio | rs274567 | -0.58 | -0.68 | -0.48 | 8.97×10^-30^ | - |
| *Whole body fat-free mass | Inverse variance weighted | 2 | -0.12 | -0.20 | -0.05 | 0.001 | 2.75×10^-3^ |
| Whole body fat-free mass | Wald ratio | rs1171614 | -0.10 | -0.13 | -0.06 | 2.31×10^-9^ | - |
| Whole body fat-free mass | Wald ratio | rs274567 | -0.18 | -0.22 | -0.13 | 9.53×10^-16^ | - |
| Whole body fat mass | Inverse variance weighted | 2 | -0.04 | -0.08 | -0.003 | 0.04 | 0.81 |

LCI = lower 95% confidence interval; UCI = upper 95% confidence interval; P_assoc_ = P-value for association; P_het_ = P-value for heterogeneity between instrumental SNP causal estimates; # SNPs = total number of SNPs used in the analysis

*These outcomes showed evidence of heterogeneity between SNPs (Phet ≤ 0.05) therefore results of single SNP MR analyses are also presented.

**Table S8. Result of our main and sensitivity MR analyses of carnitine on Covid19 related comorbidities (binary outcomes).**

| **Outcome** | **Method** | **# SNPs** | **OR** | **LCI** | **UCI** | **P_assoc_** | **P_het_** | **P_pltr_** |
| --- | --- | --- | --- | --- | --- | --- | --- | --- |
| Alzheimer's | Inverse variance weighted | 17 | 1.02 | 0.97 | 1.06 | 0.45 | 0.34 | 0.48 |
| Alzheimer's | MR Egger | 17 | 1.05 | 0.96 | 1.14 | 0.33 | - | - |
| Alzheimer's | Weighted median | 17 | 1.04 | 0.98 | 1.10 | 0.21 | - | - |
| Alzheimer's | Weighted mode | 17 | 1.06 | 1.00 | 1.12 | 0.09 | - | - |
| Breast cancer | *Inverse variance weighted | 15 | 1.09 | 0.95 | 1.26 | 0.23 | - | - |
| Breast cancer | Inverse variance weighted | 16 | 1.02 | 0.85 | 1.23 | 0.81 | 0.007 | 0.423 |
| Breast cancer | MR Egger | 16 | 1.16 | 0.82 | 1.63 | 0.42 | - | - |
| Breast cancer | Weighted median | 16 | 1.12 | 0.94 | 1.32 | 0.20 | - | - |
| Breast cancer | Weighted mode | 16 | 1.14 | 0.97 | 1.34 | 0.18 | - | - |
| Coronary heart disease | Inverse variance weighted | 17 | 1.43 | 1.07 | 1.90 | 0.01 | 0.68 | 0.17 |
| Coronary heart disease | MR Egger | 17 | 0.99 | 0.56 | 1.76 | 0.98 | - | - |
| Coronary heart disease | Weighted median | 17 | 1.14 | 0.77 | 1.69 | 0.50 | - | - |
| Coronary heart disease | Weighted mode | 17 | 1.16 | 0.73 | 1.82 | 0.50 | - | - |
| Chronic kidney disease | Inverse variance weighted | 17 | 0.95 | 0.72 | 1.27 | 0.75 | 0.50 | 0.26 |
| Chronic kidney disease | MR Egger | 17 | 1.25 | 0.73 | 2.14 | 0.42 | - | - |
| Chronic kidney disease | Weighted median | 17 | 1.13 | 0.76 | 1.67 | 0.54 | - | - |
| Chronic kidney disease | Weighted mode | 17 | 1.12 | 0.74 | 1.70 | 0.57 | - | - |
| Muscle weakness | *Inverse variance weighted | 15 | 0.93 | 0.79 | 1.10 | 0.43 | - | - |
| Muscle weakness | Inverse variance weighted | 17 | 1.04 | 0.83 | 1.32 | 0.71 | 0.0003 | 0.22 |
| Muscle weakness | MR Egger | 17 | 0.83 | 0.54 | 1.27 | 0.39 | - | - |
| Muscle weakness | Weighted median | 17 | 0.92 | 0.76 | 1.12 | 0.37 | - | - |
| Muscle weakness | Weighted mode | 17 | 0.88 | 0.72 | 1.09 | 0.23 | - | - |
| Prostate cancer | Inverse variance weighted | 17 | 1.03 | 0.83 | 1.29 | 0.78 | 0.02 | 0.77 |
| Prostate cancer | MR Egger | 17 | 1.09 | 0.71 | 1.68 | 0.70 | - | - |
| Prostate cancer | Weighted median | 17 | 1.11 | 0.88 | 1.40 | 0.33 | - | - |
| Prostate cancer | Weighted mode | 17 | 1.13 | 0.90 | 1.42 | 0.28 | - | - |
| Stroke | *Inverse variance weighted | 16 | 0.99 | 0.81 | 1.21 | 0.90 | - | - |
| Stroke | Inverse variance weighted | 17 | 1.05 | 0.81 | 1.36 | 0.74 | 4.78×10^-3^ | 0.97 |
| Stroke | MR Egger | 17 | 1.04 | 0.63 | 1.72 | 0.89 | - | - |
| Stroke | Weighted median | 17 | 1.06 | 0.83 | 1.34 | 0.66 | - | - |
| Stroke | Weighted mode | 17 | 1.05 | 0.82 | 1.35 | 0.68 | - | - |
| Type II Diabetes | *Inverse variance weighted | 15 | 1.24 | 1.02 | 1.51 | 0.05 | - | - |
| Type II Diabetes | Inverse variance weighted | 17 | 1.35 | 1.04 | 1.75 | 0.02 | 1.93×10^-5^ | 0.43 |
| Type II Diabetes | MR Egger | 17 | 1.15 | 0.71 | 1.85 | 0.58 | - | - |
| Type II Diabetes | Weighted median | 17 | 1.25 | 1.03 | 1.51 | 0.02 | - | - |
| Type II Diabetes | Weighted mode | 17 | 1.24 | 1.03 | 1.51 | 0.04 | - | - |

OR = odds ratio; LCI = lower 95% confidence interval; UCI = upper 95% confidence interval; P_assoc_ = P-value for association; P_het_ = P-value for heterogeneity between instrumental SNP causal estimates; P_pltr_ = P-value for horizontal pleiotropy from MR-Egger intercept test

\# SNPs = total number of SNPs used in the analysis

* MR-PRESSO detected outlying SNPs for these outcomes and the results were generated using inverse variance weighted method with outliers removed.

**Table S9. Results of our main and sensitivity MR analyses of carnitine on Covid19 related comorbidities (continuous outcomes)**

| **Outcome** | **Method** | **# SNPs** | **Beta** | **LCI** | **UCI** | **P_assoc_** | **P_het_** | **P_pltr_** |
| --- | --- | --- | --- | --- | --- | --- | --- | --- |
| Body Mass Index | *Inverse variance weighted | 12 | -0.02 | -0.06 | 0.04 | 0.56 | - | - |
| Body Mass Index | Inverse variance weighted | 17 | 0.007 | -0.08 | 0.09 | 0.86 | 8.23×10^-16^ | 0.64 |
| Body Mass Index | MR Egger | 17 | 0.04 | -0.12 | 0.20 | 0.63 | - | - |
| Body Mass Index | Weighted median | 17 | 0.007 | -0.04 | 0.05 | 0.75 | - | - |
| Body Mass Index | Weighted mode | 17 | 0.0002 | -0.05 | 0.05 | 0.99 | - | - |
| C-reactive protein | *Inverse variance weighted | 15 | 0.04 | -0.06 | 0.13 | 0.44 | - | - |
| C-reactive protein | Inverse variance weighted | 17 | 0.03 | -0.10 | 0.16 | 0.62 | 6.80×10^-06^ | 0.83 |
| C-reactive protein | MR Egger | 17 | 0.06 | -0.21 | 0.32 | 0.67 | - | - |
| C-reactive protein | Weighted median | 17 | 0.04 | -0.06 | 0.14 | 0.45 | - | - |
| C-reactive protein | Weighted mode | 17 | 0.06 | -0.05 | 0.16 | 0.31 | - | - |
| Height | *Inverse variance weighted | 11 | -0.17 | -0.28 | -0.06 | 0.01 | - | - |
| Height | Inverse variance weighted | 17 | -0.20 | -0.43 | 0.03 | 0.08 | 7.11×10^-45^ | 0.51 |
| Height | MR Egger | 17 | -0.34 | -0.79 | 0.12 | 0.16 | - | - |
| Height | Weighted median | 17 | -0.19 | -0.28 | -0.09 | 9.67×10^-5^ | - | - |
| Height | Weighted mode | 17 | -0.19 | -0.28 | -0.096 | 9.96×10^-4^ | - | - |
| Whole body fat mass | *Inverse variance weighted | 15 | -0.08 | -0.12 | -0.03 | 0.24 | - | - |
| Whole body fat mass | Inverse variance weighted | 17 | -0.04 | -0.11 | 0.03 | 0.22 | 3.36×10^-6^ | 0.97 |
| Whole body fat mass | MR Egger | 17 | -0.04 | -0.18 | 0.09 | 0.56 | - | - |
| Whole body fat mass | Weighted median | 17 | -0.05 | -0.10 | 0.005 | 0.08 | - | - |
| Whole body fat mass | Weighted mode | 17 | -0.05 | -0.10 | -0.002 | 0.06 | - | - |
| Whole body fat-free mass | *Inverse variance weighted | 10 | -0.07 | -0.10 | -0.04 | 0.002 | - | - |
| Whole body fat-free mass | Inverse variance weighted | 17 | -0.07 | -0.14 | 0.009 | 0.09 | 2.30×10^-25^ | 0.47 |
| Whole body fat-free mass | MR Egger | 17 | -0.11 | -0.25 | 0.03 | 0.15 | - | - |
| Whole body fat-free mass | Weighted median | 17 | -0.07 | -0.11 | -0.04 | 3.69×10^-5^ | - | - |
| Whole body fat-free mass | Weighted mode | 17 | -0.08 | -0.11 | -0.04 | 3.92×10^-4^ | - | - |

LCI = lower 95% confidence interval; UCI = upper 95% confidence interval; P_assoc_ = P-value for association; P_het_ = P-value for heterogeneity between instrumental SNP causal estimates; P_pltr_ = P-value for horizontal pleiotropy from MR-Egger intercept test

\# SNPs = total number of SNPs used in the analysis

* MR-PRESSO detected outlying SNPs for these outcomes and the results were generated using inverse variance weighted method with outliers removed.

**Table S10. Multivariable MR results**

| **Exposure** | **Outcome** | **Adjusted for** | **#SNP** | **OR/Beta** | **LCI** | **UCI** | **P** |
| --- | --- | --- | --- | --- | --- | --- | --- |
| **Acetyl-carnitine** | | | | | | | |
|  | Severe Covid19 | Body Mass Index | 2 | 0.69 | 0.45 | 1.05 | 0.08 |
|  | Severe Covid19 | Coronary heart disease | 2 | 0.67 | 0.36 | 1.27 | 0.22 |
|  | Severe Covid19 | Type II Diabetes | 1 | 0.77 | 0.47 | 1.25 | 0.29 |
|  | C-reactive protein | Body Mass Index | 1 | -0.14* | -0.25 | -0.04 | 0.007 |
| **Carnitine** | | | | | | | |
|  | Severe Covid19 | Body Mass Index | 14 | 0.63 | 0.38 | 1.05 | 0.08 |
|  | Severe Covid19 | Coronary heart disease | 17 | 0.58 | 0.33 | 1.04 | 0.07 |
|  | Severe Covid19 | Type II Diabetes | 14 | 0.64 | 0.39 | 1.05 | 0.08 |
|  | C-reactive protein | Body Mass Index | 14 | 0.47* | -0.68 | 1.61 | 0.42 |

LCI = lower 95% confidence interval; UCI = upper 95% confidence interval; P = P-value for association; # SNPs = total number of SNPs used in the analysis

* effect estimates (betas) are given for continuous outcomes.

| **A)**  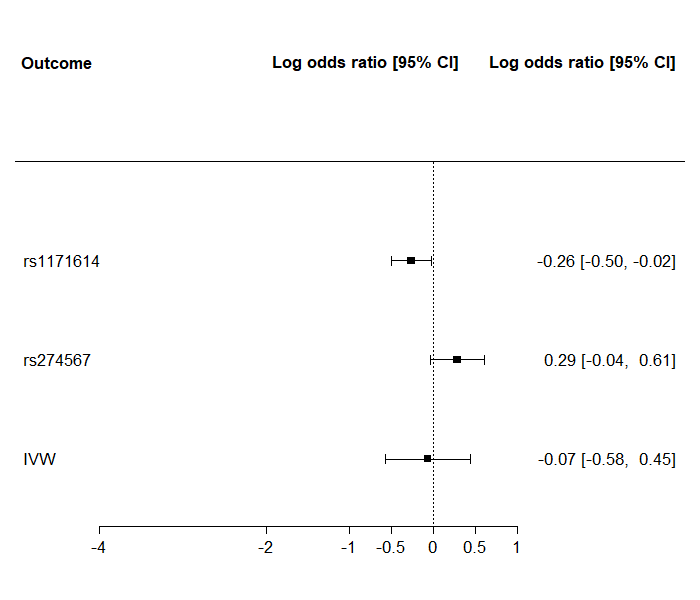 | **B)**  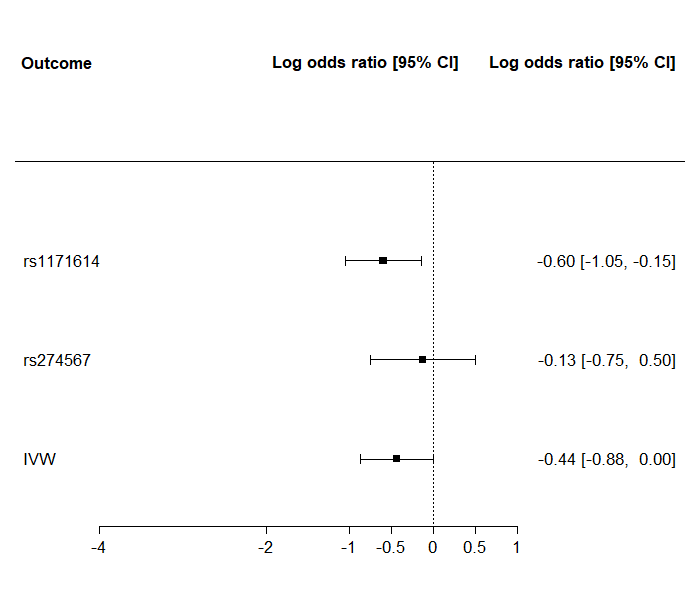 |
| --- | --- |
| **C)**  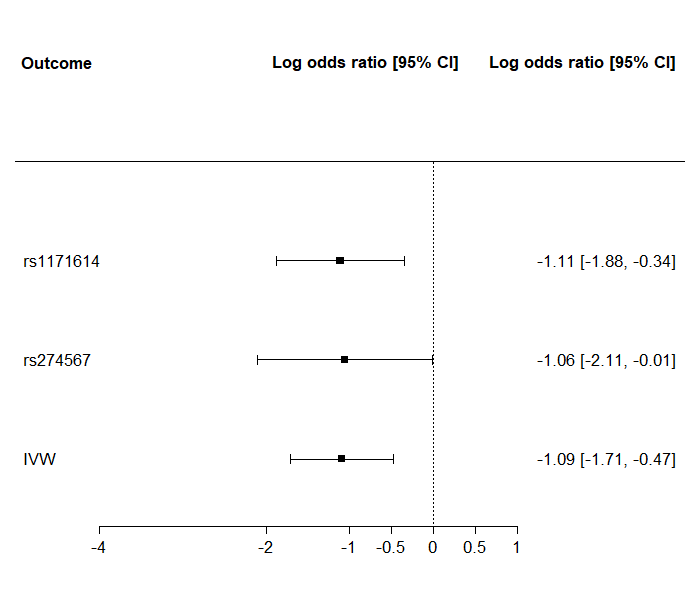 | **D)**  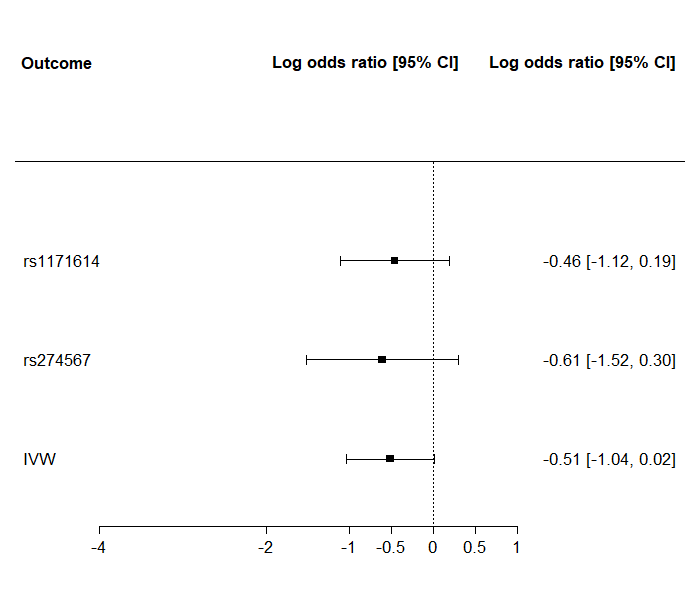 |

**Figure S1. Forest plot showing odds ratio estimates of acetyl-carnitine on A) any Covid19; B) hospitalisation with Covid19 vs population controls; C) hospitalisation with Covid19 versus controls with Covid19 infection; D) severe Covid19 versus population controls for each of the SNPs for acetyl-carnitine individually and combined.**

**Figure S2. Forest plot showing odds ratio estimates of carnitine on A) any Covid19; B) hospitalisation with Covid19 vs population controls; C) hospitalisation with Covid19 versus controls with Covid19 infection; D) severe Covid19 versus population controls for each of the SNPs for acetyl-carnitine individually and combined.**

| **A)** 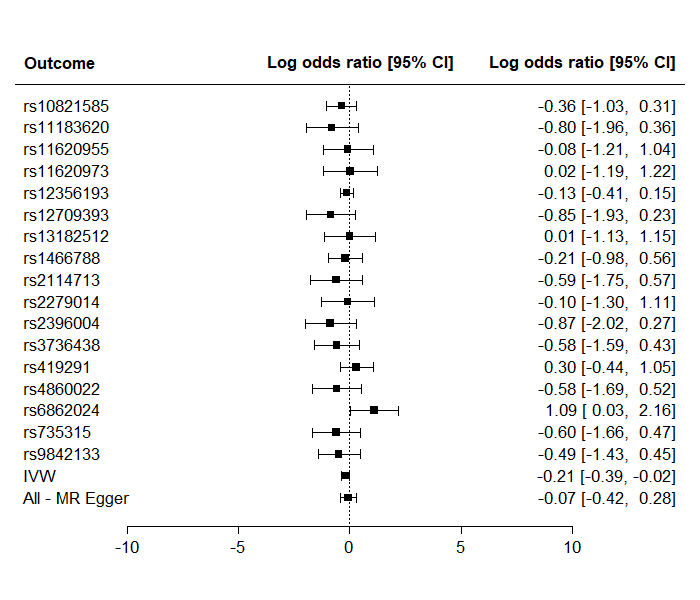 | **B)** 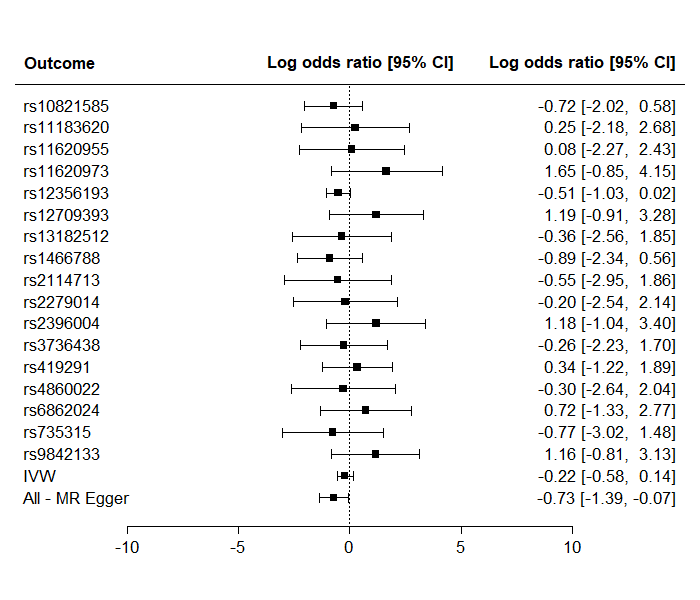 |
| --- | --- |
| **C)**  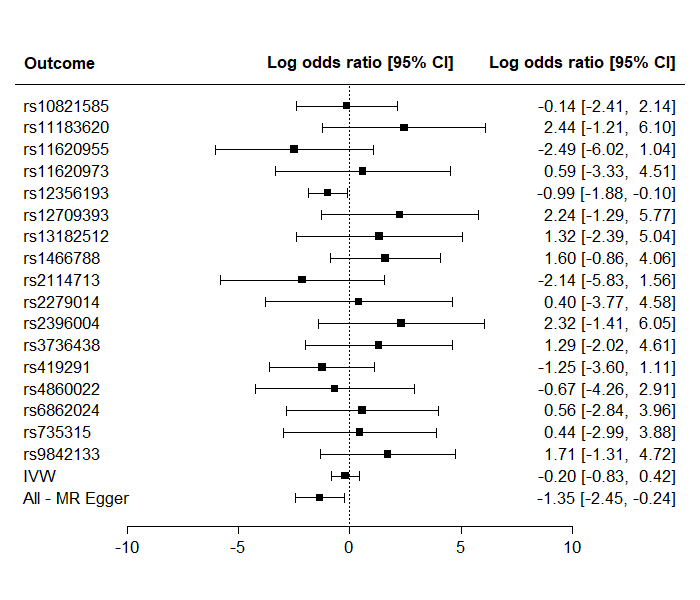 | **D)**  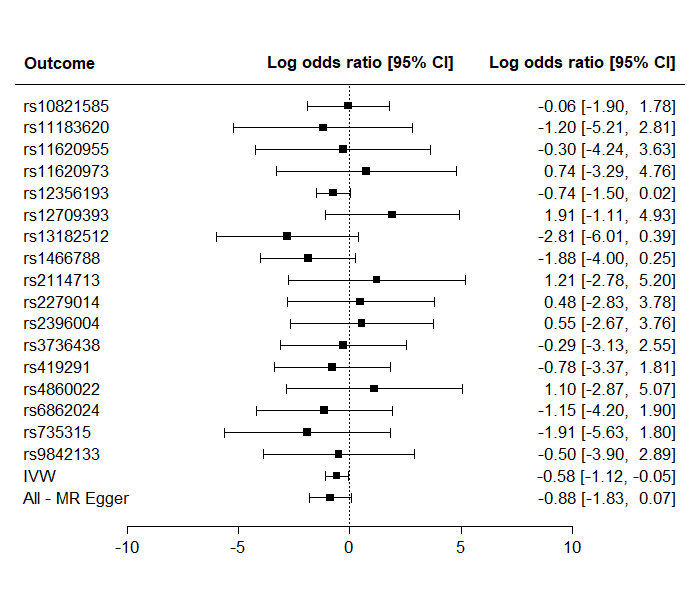 |

**Figure S3. Comparison of results using different MR methods for carnitine and A) any Covid19; B) hospitalisation with Covid19 vs population controls; C) hospitalisation with Covid19 versus controls with Covid19 infection; D) severe Covid19 versus population controls.**

| **A)**  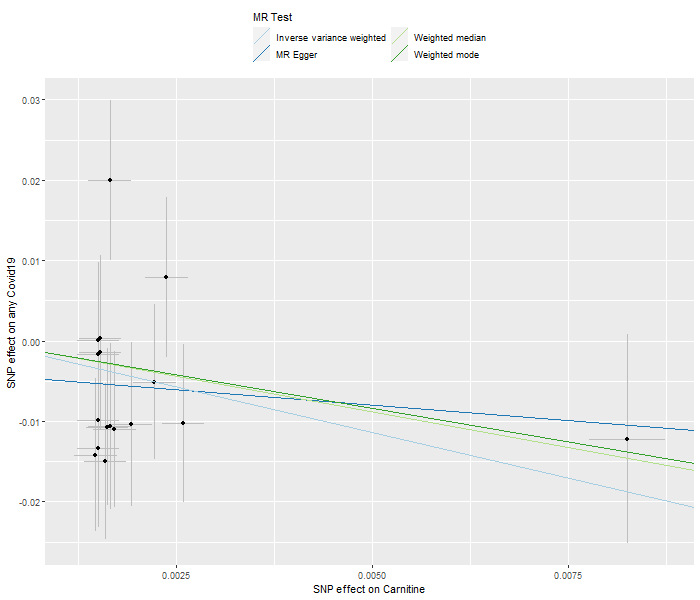 | **B)**  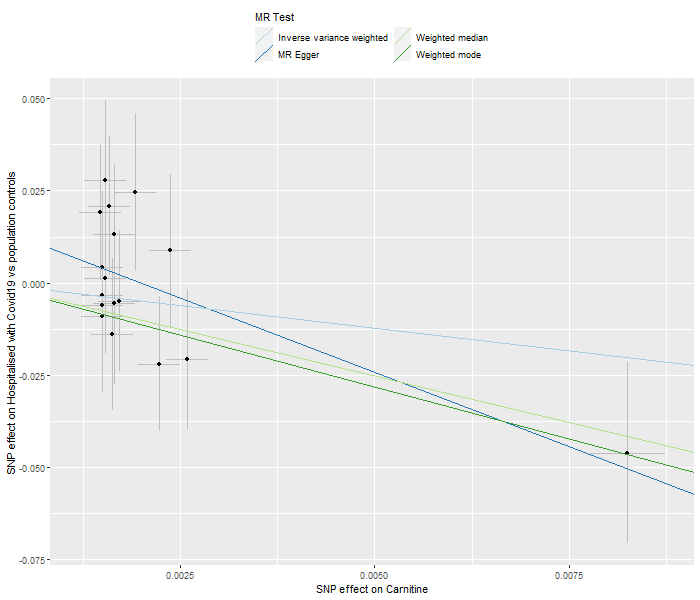 |
| --- | --- |
| **C)** 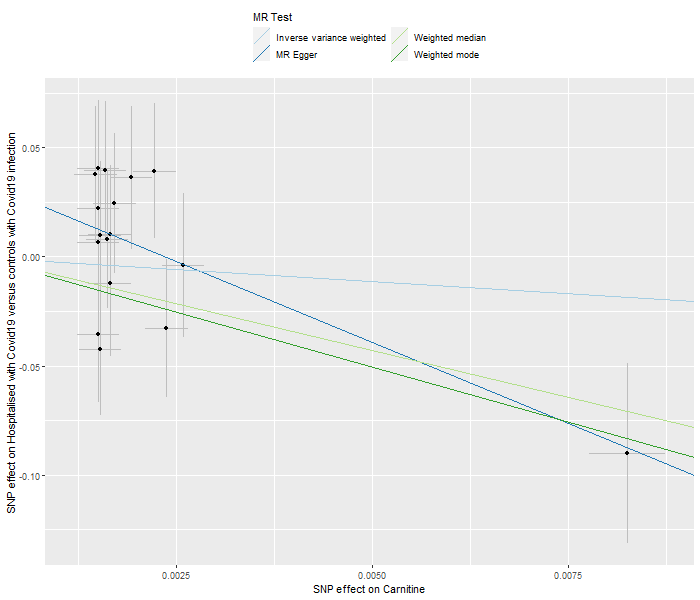 | **D)** 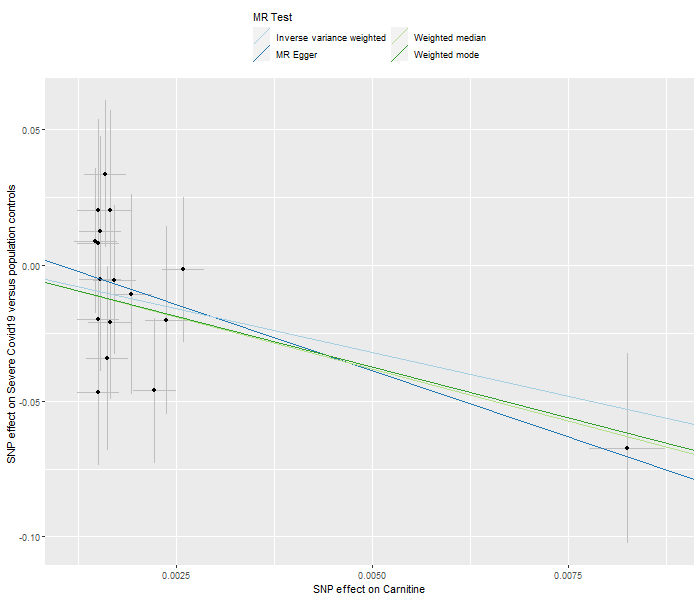 |

**Figure S4. MR estimates for acetyl-carnitine concentration on Covid19 related comorbidities**

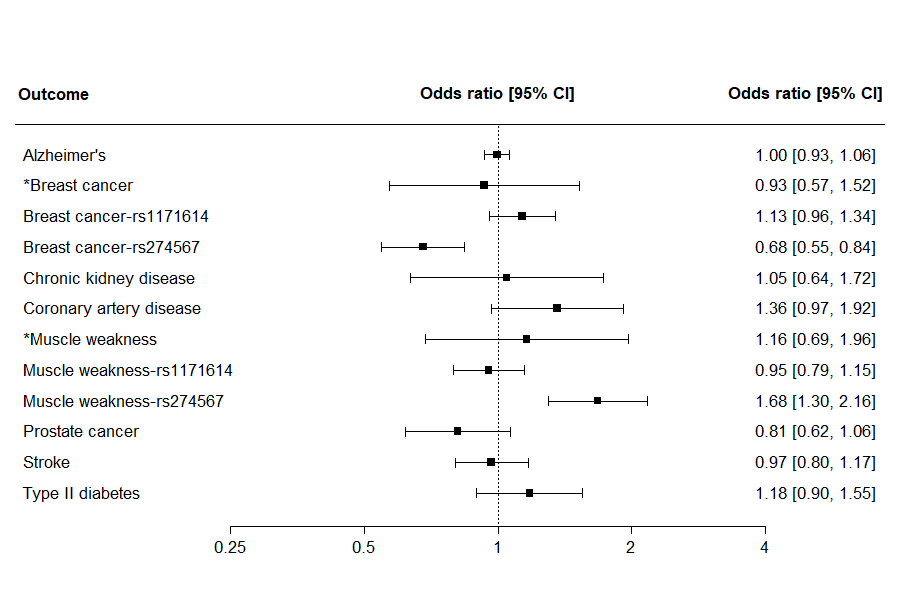

*These outcomes showed evidence of heterogeneity between SNPs (P_het_ ≤ 0.05) therefore we presented MR results using both IVW and Wald ratio for each SNPs.

**Figure S5. MR estimates for carnitine concentration on Covid19 related comorbidities**

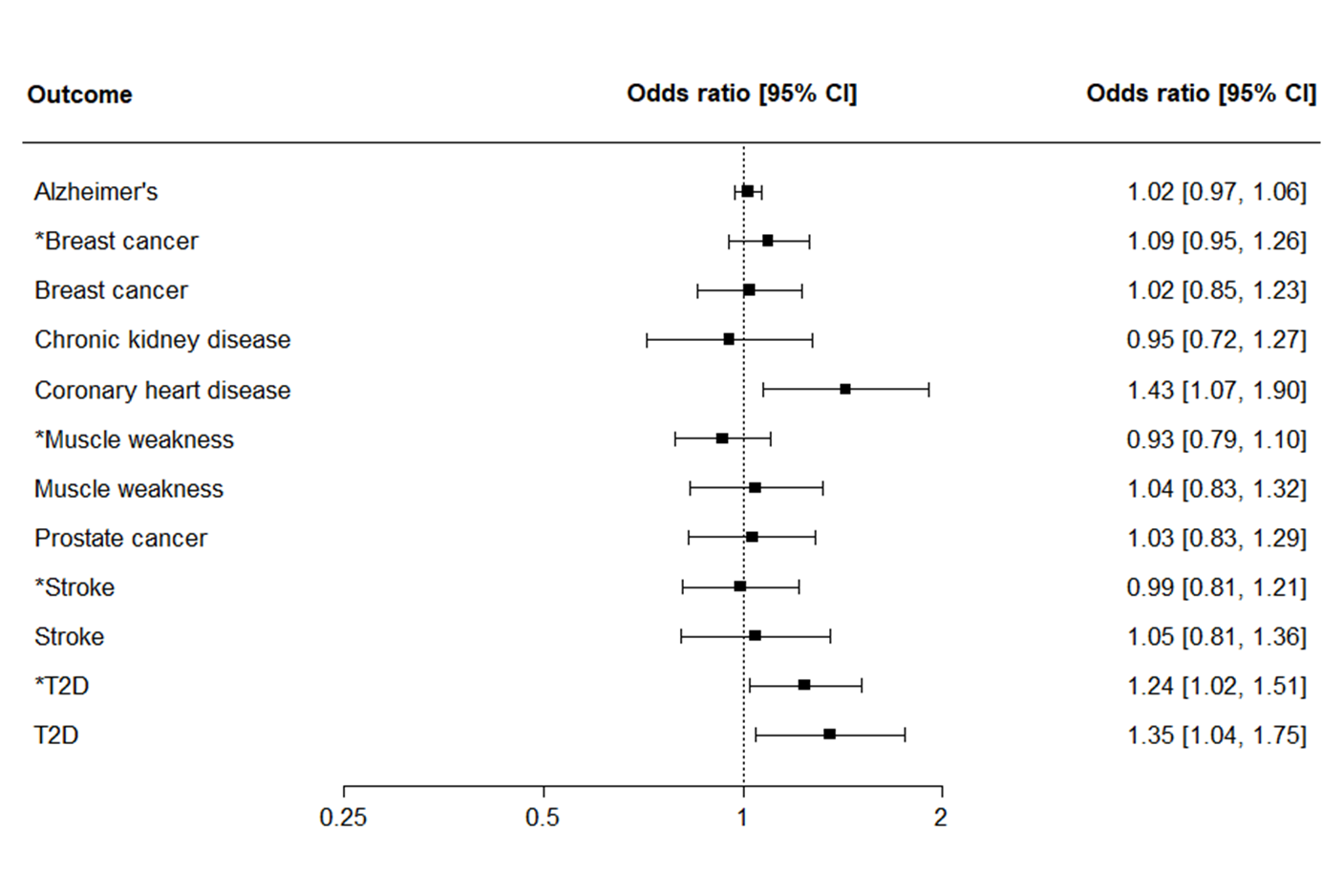

*These outcomes showed evidence of heterogeneity between SNPs (Phet ≤ 0.05) and MR-PRESSO detected outlying SNPs for these outcomes these results were generated using inverse variance weighted method with outliers removed.
